# Prospective Clinical Surveillance for Severe Acute Respiratory Illness in Kenyan Hospitals during the COVID-19 pandemic

**DOI:** 10.1101/2024.03.03.24303690

**Authors:** RK Lucinde, H Gathuri, L Isaaka, M Ogero, L Mumelo, D Kimego, G Mbevi, C Wanyama, EO Otieno, S Mwakio, M Saisi, E Isinde, IN Oginga, A Wachira, E Manuthu, H Kariuki, J Nyikuli, C Wekesa, A Otedo, H Bosire, SB Okoth, W Ongalo, D Mukabi, W Lusamba, B Muthui, I Adembesa, C Mithi, M Sood, N Ahmed, B Gituma, M Giabe, C Omondi, R Aman, P Amoth, K Kasera, F Were, W Nganga, JA Berkley, BA Tsofa, J Mwangangi, P Bejon, E Barasa, M English, JAG Scott, S Akech, EW Kagucia, A Agweyu, AO Etyang

**Author notes:** Corresponding author: Ruth K. Lucinde KEMRI-Wellcome Trust Research Programme (KWTRP), Nairobi, Kenya.

## Abstract

**Background:** There are limited data from sub-Saharan Africa describing the pattern of admissions to public hospitals with severe acute respiratory infections during the COVID-19 pandemic.

**Methods:** We conducted a prospective longitudinal hospital-based sentinel surveillance between May 2020 and December 2022 at 16 public hospitals in Kenya. All patients aged above 18 years admitted to adult medical wards in the participating hospitals were included. Demographic and clinical characteristics, COVID-19 infection and vaccination status and outcome data were collected.

**Results:** Of the 52,714 patients included in the study, 18,001 (35%) were admitted with severe acute respiratory illness (SARI). The mean age was 51 years. Patients were equally distributed across sexes. Pneumonia was the most common diagnosis at discharge. Hypertension, HIV and diabetes mellitus were the most common comorbidities. COVID-19 test results were positive in 2,370 (28%) of the 8,517 (47%) patients that underwent testing.

Overall in-patient case fatality for SARI was 21% (n=3,828). After adjusting for age, sex and presence of a comorbidity, SARI patients had higher in-patient mortality compared to non-SARI patients regardless of their COVID-19 status (aHR 1.31, 95% CI 1.19 – 1.46). COVID-19 positive SARI patients had a higher in-patient mortality rate compared to their negative counterparts (aHR 1.31, 95% CI 1.12 - 1.54, p value < 0.0001).

COVID-19 vaccine effectiveness against mortality due to SARI after adjusting for age, sex and presence of a comorbidity was 34% (95% CI 11% - 51%).

**Discussion:** We have provided a comprehensive description of the pattern of admissions with respiratory illnesses in Kenyan hospitals during the COVID-19 pandemic period. We have demonstrated the utility of routine surveillance activities within public hospitals in low-income settings which if strengthened can enhance the response to emerging health threats.

## Background

Unexpected disease outbreaks such as the COVID-19 pandemic pose unique challenges in characterising the clinical pattern of disease due to their novelty as well as the evolving patterns of clinical manifestation based on pathogen strains(1). By 5^th^ May 2023 when the COVID-19 pandemic was declared over by the World Health Organisation (WHO), Kenya had reported more than 340,000 confirmed cases and over 5,000 deaths due to COVID-19 against a global tally of over 770 million cases and almost 7 million deaths(2). Three years into the pandemic, the pattern of disease among patients admitted to hospital with the disease is still being described with key differences in severity and outcome by specific groups such as age, comorbidities or ethnic groups being apparent(3, 4, 5, 6, 7). However, data on these patterns in Kenyan and other sub-Saharan populations is limited, despite indications that the clinical pattern of illness in these settings was significantly different to that in high-income countries (8).

Globally, hospital-based surveillance systems have been key in generating timely data on the evolving clinical pattern of COVID-19 and informing clinical practice(4, 5, 9, 10, 11, 12). These systems have been key in evaluating trends in mortality, admissions and disease burden as well as evaluating vaccine effectiveness against severe disease and mortality (9).

Kenya’s national COVID-19 vaccination programme begun on 5^th^ March 2021. As at December 2023, over 23 million vaccine doses of six licensed vaccine products (Moderna, Pfizer/BioNTech, Gamaleya, Janssen(Johnson & Johnson), Oxford/AstraZeneca and Sinopharm) (13) have been delivered (2). However, vaccine coverage in the country remains low at about 36% among adults (2, 14) while no local data on the effectiveness of these vaccines exist.

Working with Ministry of Health (MOH) officials in Kenya, we developed a hospital-based surveillance system using existing infrastructure from a clinical information network (CIN)(15, 16, 17) to conduct surveillance for admissions with severe acute respiratory illness (SARI) and COVID-19 in Kenya. We used this platform to evaluate the effectiveness of licensed COVID-19 vaccines against admission with severe acute respiratory disease in Kenya.

This study describes the characteristics of adults admitted to the medical wards of 16 public hospitals with Severe Acute Respiratory Illness (SARI) in Kenya between 5^th^ May 2020 and 31^st^ December 2022. We explore risk factors for poor outcomes overall and among those who tested positive for COVID-19. Additionally, we describe the effectiveness of licensed COVID-19 vaccines against admission with COVID-19 as well as mortality due to SARI.

## Methods

### Study Design

We conducted a prospective multi-hospital clinical surveillance between 5^th^ May 2020 and 31^st^ December 2022 in 16 public hospitals in Kenya (Supplementary Figure 1). The study sites were level four and level five hospitals part of a clinical information network (CIN) established in 2013(17). They are spread across the country’s densely populated western, central and coastal regions(18).

The surveillance was aimed at monitoring the evolution of the pandemic through review of admissions and management of patients with SARI in the adult medical wards. Additionally, it allowed for the estimation of COVID-19 vaccine effectiveness using a test-negative case-control study approach. Data were collected conducted during the course of routine delivery of care and we did not influence the clinical management of patients in the wards or provide any support for laboratory testing of COVID-19.

This study was approved by the Scientific and Ethical Review Unit (SERU) of the Kenya Medical Research Institute (KEMRI).

### Study Patients

Data from all adults (age ≥18 years) patients discharged from medical wards in the participating sites during the study period were included. The discharge date and diagnosis were used to define in-patient episodes to ensure consistency in populations(18).

We collected data on the demographic and clinical characteristics, COVID-19 vaccination status, discharge diagnoses and clinical outcome of all included patients. Those with SARI had additional data on their clinical symptoms, laboratory results and management collected.

Using Kenyan MOH guidelines, SARI was defined as the presence of fever, cough or shortness of breath that required hospitalisation, in the absence of an alternative diagnosis that fully explains the clinical presentation(19, 20). Confirmed COVID-19 cases were those that had a positive real-time reverse transcription-polymerase chain reaction (RT-PCR) or antigen detection rapid diagnostic test (Ag-RDT) for severe acute respiratory syndrome coronavirus 2 (SARS-CoV-2) in a respiratory sample during or before admission(20, 21).

Admission and discharge diagnoses were recorded based on the International Classification of Diseases, Tenth Revision, Clinical Modification (ICD-10-CM) codes.

### Data Collection

The data collection and analysis procedures for the CIN sites are reported elsewhere(22). In summary, trained clinical officers abstracted data from patient records to an electronic data capture tool at each site daily. De-identified data were then synchronized to KEMRI-Wellcome Trust Research Programme (KWTRP) servers where a central data team conducted regular data checks. Additional checks included cross-validation of monthly tallies with daily bed returns reported at the hospitals(18).

A dynamic structured data collection tool was adapted for use. The tool was updated regularly in-keeping with emerging knowledge and changes in global and national guidelines for COVID-19 patient management. Key changes included a.) recording of steroid use among patients admitted with SARI following the WHO recommendation of dexamethasone use in management of COVID-19 in June 2020(20, 23); b.) use of Ag-RDT results in addition to RT-PCR results to confirm positive COVID-19 cases(21) and, c.) recording of patient vaccination status following the commencement of the COVID-19 vaccination programme in Kenya in March 2021(24).

### Statistical Analysis

#### Mortality among patients admitted to the adult medical wards

We described the characteristics of all patients in the study. We determined factors associated with mortality during the hospital admission using a forward-built stepwise Cox regression model. Patients entered the risk set at admission and exited at the earliest of either date of death or date of discharge from hospital. The final model included age, sex and presence of a comorbidity as risk factors for mortality.

#### Predictors of mortality among patients with SARI and patients with COVID-19

We used a forward stepwise approach to build a Cox regression model to determine the factors associated with death among SARI patients in general and among SARI patients who tested positive for COVID-19. This analysis was limited to patients who had complete data on all potential risk factors, including age, sex, presence of a comorbidity, leucocyte count, haemoglobin level, temperature oxygen saturation at admission and COVID-19 test results. These variables were defined as confounders if the crude and adjusted rate ratio differed by >10% and/or the stratum specific rate ratios differed by >10%.

#### COVID-19 Vaccine Effectiveness

We used a test-negative case control design to estimate COVID-19 vaccine effectiveness (VE). We restricted these analyses to patients admitted after 5^th^ March 2021 when COVID-19 vaccines were available in Kenya.

Patients were considered vaccinated if they had received a COVID-19 vaccine at least 14 days before their admission. We used logistic regression, to estimate COVID-19 VE against the following outcomes: (a) admission with COVID-19, and (b) mortality due to SARI (disregarding COVID-19 status because of low COVID-19 testing at the hospitals). We in addition computed VE estimates by (a) vaccine product and (b) the predominant circulating SARS-CoV-2 variant.

For VE against admission with COVID-19, we considered as cases SARI patients who tested positive for COVID-19 while those who tested negative were considered controls. For VE against mortality due to SARI, we considered SARI patients who died during admission as cases while those who were discharged alive were considered controls. Estimates were adjusted by age, sex and presence of a comorbidity. VE was calculated as 1 minus the adjusted odds ratio multiplied by 100.

We could not estimate VE by full/partial vaccination status and VE against mortality due to COVID-19 because of limited COVID-19 testing, incomplete vaccination records, difficulty confirming the vaccination status for some participants from the national vaccination registry and mixing of vaccine products.

All data were analysed using Stata/IC™ 16.0 (Stata Corp, College Station, Texas, USA).

## Results

### General characteristics of study patients

We collected data from 52,714 patients admitted across the 16 hospitals. Of these, 18,001(34.2%) were patients classified as having SARI. Of all SARI patients, 4,883(27.1%) had complete data for inclusion in the analysis for predictors of mortality among SARI patients.

The mean age of patients was 51 years (Standard deviation (SD) ±8 20). Distribution of patients by sex was equal. The median length of stay in hospital was 5 days (Interquartile range (IQR) 3 to 9 days).

Pneumonia was the most common discharge diagnosis with heart failure and pulmonary tuberculosis being the second and third, respectively. One or more comorbidities was present at admission in 16.5% (n=8,683) of all patients. Hypertension (6.5%, n= 3,755) was the most common comorbidity. HIV infection was present in 2,690 patients (5.1%), while 1,975(3.8%) of all patients had Diabetes Mellitus.

The proportion of patients receiving specialised care as part of their management i.e., receiving ventilation, IV fluids and/or oxygen was highest during the second quarter of 2021 (13.1%; n=1,342) and lowest during the last quarter of 2022 (2.8%; n=284). Overall, 16.8% (n=8,848/52,714) of the included patients died in hospital (Table 1).

**Table 1:** Distribution of patients by key demographic and clinical parameters

| Category<br>(*n: all, SARI) |  | All patients<br>n (%) | SARI patients<br>n (%) |
| --- | --- | --- | --- |
| Age |  |  |  |
| (52,711; 18,000) | 18 – 30 years | 8,369 (15.9%) | 2,020 (11.2%) |
|  | 31 – 40 years | 9,292 (17.6%) | 2,952 (16.4%) |
|  | 41 – 50 years | 8,465 (16.1%) | 2,799 (15.6%) |
|  | 51 – 60 years | 7,784 (14.8%) | 2,965 (16.5%) |
|  | 61 years and above | 18,801 (35.75%) | 7,264 (40.4%) |
| Sex |  |  |  |
| (52,703; 17,995) | Female | 26,453 (50.2%) | 8,492 (47.2%) |
|  | Male | 26,250 (49.8%) | 9,503 (52.8%) |
| Presence of comorbidity |  |  |  |
| (52,714; 18,001) | None | 44,031 (83.5%) | 9,404 (52.2%) |
|  | One | 7,048 (13.4%) | 6,978 (38.8%) |
|  | Two | 1,512 (2.9%) | 1,496 (8.3%) |
|  | Three or more | 123 (0.23%) | 123 (0.7%) |
| Length of stay |  |  |  |
| (52,616; 17,978) | Less than 5 days | 22,632 (43.0%) | 6,792 (37.8%) |
|  | 5 – 7 days | 8,552 (16.3%) | 2,972 (16.5%) |
|  | 1 week – 2 weeks | 14,290 (27.2%) | 5,405 (30.1%) |
|  | More than 2 weeks | 7,142 (13.6%) | 2,809 (15.6%) |
| Outcome |  |  |  |
| (52,658; 17,983) | Dead | 8,848 (16.8%) | 3,828 (21.3%) |
|  | Alive | 43,810 (83.2%) | 14,155 (78.7%) |
| Temperature |  |  |  |
| (<sup>a</sup>, 15,837) | Hypothermia (< 36.0°C) | <sup>a</sup> | 2,342 (14.8%) |
|  | Normal | <sup>a</sup> | 11,628 (73.4%) |
|  | Hyperthermia (> 37.7 °C) | <sup>a</sup> | 1,867 (11.8%) |
| Oxygen Saturation |  |  |  |
| (<sup>a</sup>, 16,674) | Less than 90% | <sup>a</sup> | 6,519 (38.8%) |
|  | 90% to 94% | <sup>a</sup> | 4,150 (24.7%) |
|  | More than 95% | <sup>a</sup> | 6,123 (36.5%) |
| White Blood Cell count |  |  |  |
| (<sup>a</sup>, 10,979) | Leucopenia (< 4.5*10<sup>9</sup>/L) | <sup>a</sup> | 1,588 (14.3%) |
|  | Normal | <sup>a</sup> | 6,614 (59.7%) |
|  | Leukocytosis (> 12*10<sup>9</sup>/L) | <sup>a</sup> | 2,884 (26.0%) |
| Hemoglobin levels |  |  |  |
| (<sup>a</sup>, 13,841) | Anemia (< 11.5 g/dL) | <sup>a</sup> | 5,526 (39.6%) |
|  | Normal | <sup>a</sup> | 8,426 (60.4%) |
| Malaria results |  |  |  |
| (<sup>a</sup>, 4,456) | Negative | <sup>a</sup> | 3,998 (89.7%) |
|  | Positive | <sup>a</sup> | 458 (10.3%) |
| COVID results |  |  |  |
| (<sup>a</sup>, 8,517) | Negative | <sup>a</sup> | 6,147 (72.2%) |
|  | Positive | <sup>a</sup> | 2,370 (27.8%) |
| *n for each parameter varies due to variable documentation of these data |  |  |  |
| <sup>a</sup> Data not collected |  |  |  |

COVID-19 vaccination histories were available for 22,943 (43.5%) eligible patients. Of these, 4.0% (n=919) had received at least one dose of a COVID-19 vaccine (Table 1). A COVID-19 test sample was collected from 52.8% (n=8,738) of all SARI patients and results were available for 97.5% (n=8,517) of these (Table 1).

### Mortality among patients admitted to the adult medical wards

Figure 1 displays cumulative in-patient mortality for SARI and non-SARI patients. Overall, those with SARI had higher mortality rates compared to those without SARI. Case fatality ratio (CFR) for SARI patients with COVID-19 was 531/2,368 (22.4%) compared to 3,297/15,615 (21.1%) for SARI patients without COVID-19. Case fatality among patients without SARI was 14.5% (5,020/34,675). Using non-SARI patients as the reference category, the crude hazard ratio (HR) for mortality was 1.31 (95% CI 1.25 - 1.38; p value < 0.0001) for SARI patients who were negative for COVID-19. And 1.46 (95% CI 1.33 - 1.61; p value < 0.0001) for SARI patients who tested positive for COVID-19. The difference was slightly reduced after adjusting for age, sex and presence of a comorbidity (COVID-19 negative SARI aHR 1.22, 95% CI 1.15 – 1.30; COVID-19 positive SARI aHR 1.31, 95% CI 1.19 – 1.46; p-value < 0.0001). Male sex, presence of a comorbidity and increasing age were all associated with a higher rate of mortality.

**Figure 1:**
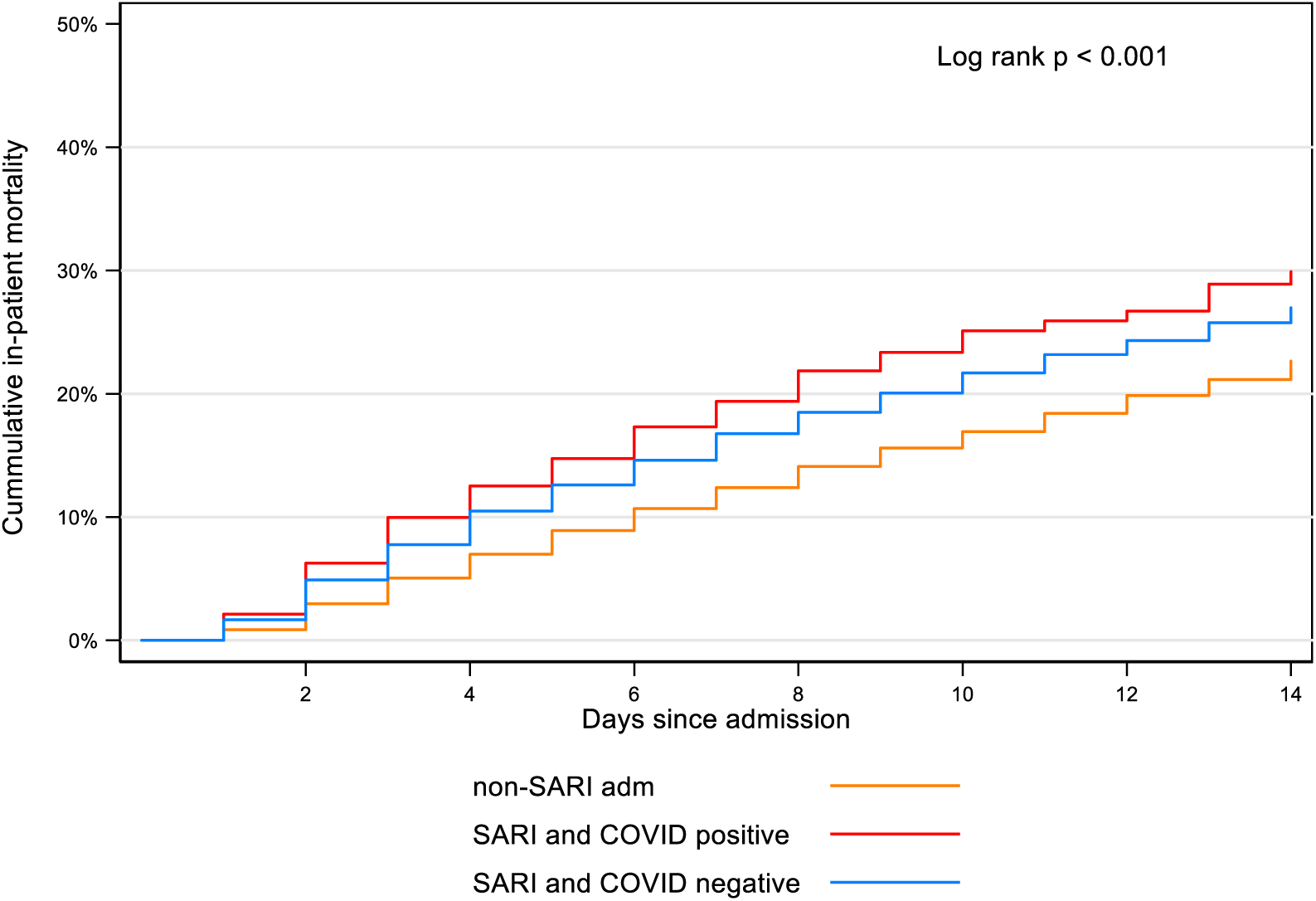
In-patient mortality among SARI and non-SARI patients

Table 2 displays the predictors of mortality due to SARI after adjusting for COVID-19 status, age group, presence of a comorbidity, temperature, oxygen saturation, white blood cell count and haemoglobin levels at admission. Overall, male sex and leucocytosis were the strongest predictors of mortality.

**Table 2:** Predictors of mortality among patients with SARI.

|  |  | **Mortality**<br>**rate per**<br>**100 person**<br>**days** | **Crude**<br>**Rate**<br>**ratio** | **95% CI** | **Adjusted**<br>**rate ratio** | **95% CI** | **LRT p**<br>**value** |
| --- | --- | --- | --- | --- | --- | --- | --- |
| COVID status | Negative | 1.5 |  |  |  |  | 0.0012 |
|  | Positive | 1.9 | **1.30** | **1.12, 1.53** | **1.31** | **1.12, 1.54** |  |
| Age | 18 – 30 years | 1.5 |  |  |  |  | 0.0049 |
|  | 31 – 40 years | 1.7 | 1.12 | 0.81, 1.53 | 1.05 | 0.77, 1.44 |  |
|  | 41 – 50 years | 1.4 | 1.02 | 0.74, 1.40 | 0.92 | 0.67, 1.27 |  |
|  | 51 – 60 years | 1.6 | 1.00 | 0.73, 1.37 | 0.90 | 0.65, 1.22 |  |
|  | 61 years and above | 1.9 | **1.38** | **1.05, 1.81** | 1.25 | 0.94, 1.66 |  |
| Sex | Female | 1.2 |  |  |  |  | 0.0001 |
|  | Male | 1.9 | **1.35** | **1.16, 1.56** | **1.37** | **1.18, 1.59** |  |
| Presence of a<br>comorbidity | None | 1.8 |  |  |  |  | 0.4891 |
|  | At least one | 1.6 | 1.02 | 0.88, 1.17 | 1.05 | 0.91, 1.22 |  |
| Temperature | Normal | 1.6 |  |  |  |  | 0.0766 |
|  | Hypothermia (< 36.0°C) | 2.0 | 1.20 | 1.00, 1.46 | **1.23** | **1.02, 1.49** |  |
|  | Hyperthermia (> 37.7 °C) | 1.8 | 1.08 | 0.86, 1.35 | 1.11 | 0.89, 1.39 |  |
| Oxygen Saturation | Less than 90% | 2.0 |  |  |  |  | < 0.0001 |
|  | 90% to 94% | 1.5 | **0.68** | **0.56, 0.83** | **0.69** | **0.57, 0.84** |  |
|  | More than 95% | 1.3 | **0.64** | **0.54, 0.75** | **0.64** | **0.54, 0.77** |  |
| White Blood Cell<br>count | Normal | 1.5 |  |  |  |  | 0.0002 |
|  | Leucopenia (< 4.5) | 1.6 | 1.06 | 0.85, 1.33 | 1.11 | 0.90, 1.39 |  |
|  | Leukocytosis (> 12) | 2.1 | **1.37** | **1.17, 1.61** | **1.38** | **1.18, 1.62** |  |
| Hemoglobin levels | Normal | 1.6 |  |  |  |  | < 0.0001 |
|  | Anemia (< 11.5) | 1.8 | 1.08 | 0.94, 1.25 | **1.26** | **1.08, 1.47** |  |

### COVID-19 Vaccine Effectiveness

Of all vaccinated participants included in the VE analysis (n =356), 20.5% (n=73) had received the AstraZeneca, 8.4%(n=30) had received the Janssen, 3.4%(n=12) had received the Moderna and 7.6%(n=27) had received the Pfizer vaccine product as their first dose. The rest, 60% (n=214) reported being vaccinated but did not have a vaccine certificate to verify the vaccine product they’d received. COVID-19 vaccine effectiveness (VE) against mortality due to SARI after adjusting for age, sex and presence of a comorbidity was 34%, 95% CI 11% - 51%, Table 3).

**Table 3:**
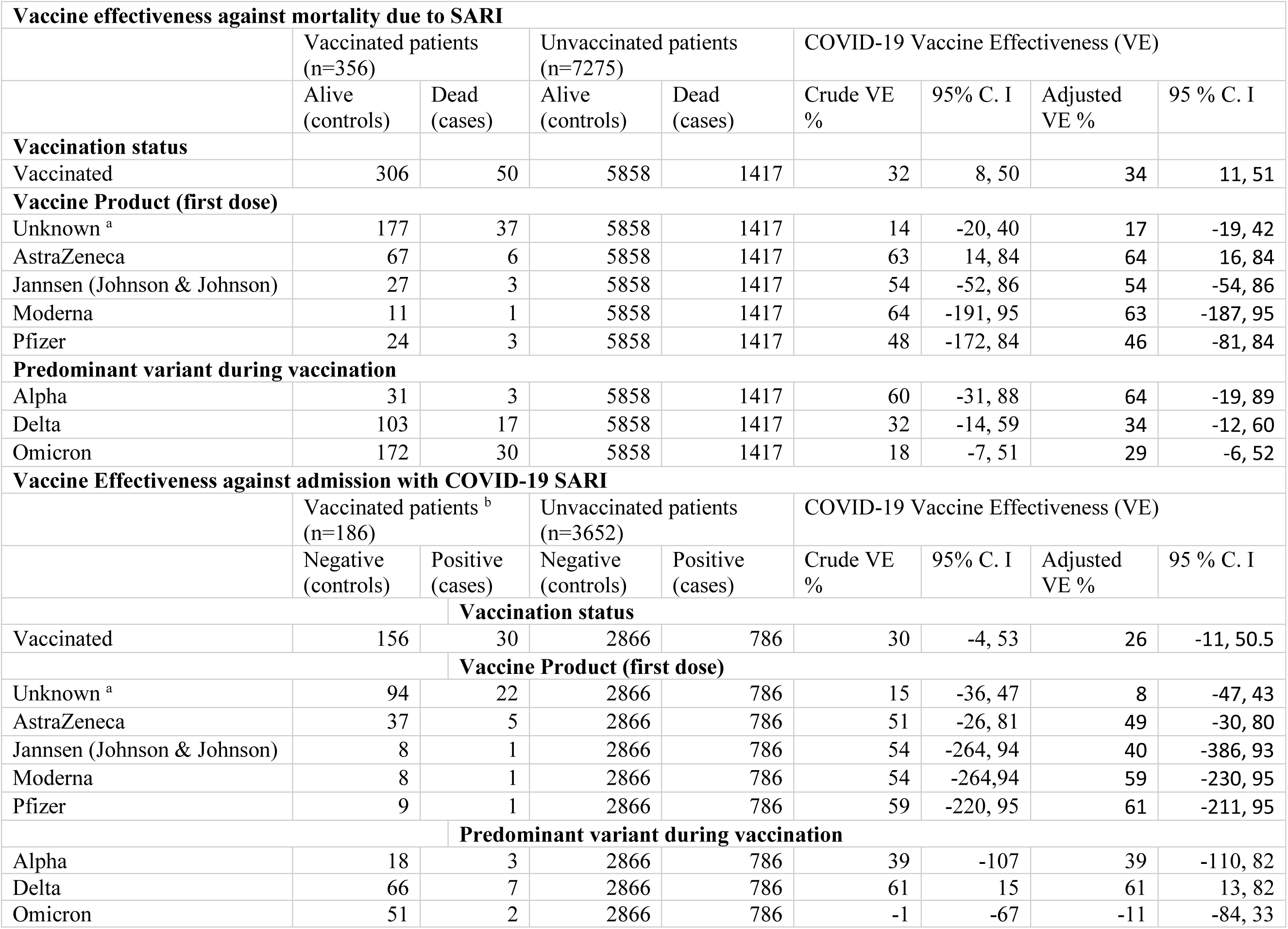
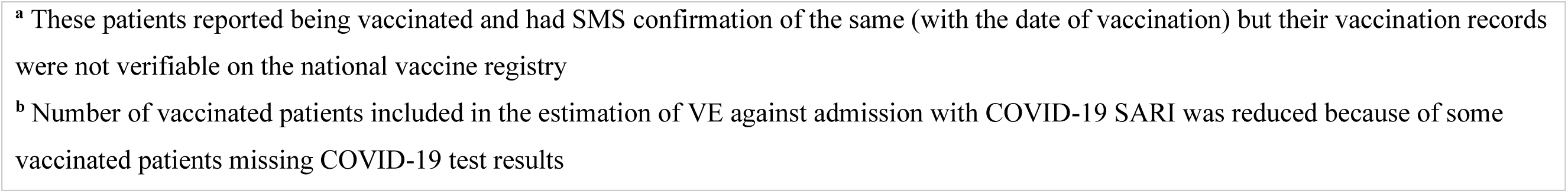
COVID-19 Vaccine Effectiveness.

| **Vaccine effectiveness against mortality due to SARI** |  |  |  |  |  |  |  |  |
| --- | --- | --- | --- | --- | --- | --- | --- | --- |
|  | Vaccinated patients<br>(n=356) |  | Unvaccinated patients<br>(n=7275) |  | COVID-19 Vaccine Effectiveness (VE) |  |  |  |
|  | Alive<br>(controls) | Dead<br>(cases) | Alive<br>(controls) | Dead<br>(cases) | Crude VE<br>% | 95% C. I | Adjusted<br>VE % | 95 % C. I |
| **Vaccination status** |  |  |  |  |  |  |  |  |
| Vaccinated | 306 | 50 | 5858 | 1417 | 32 | 8, 50 | 34 | 11, 51 |
| **Vaccine Product (first dose)** |  |  |  |  |  |  |  |  |
| Unknown <sup>a</sup> | 177 | 37 | 5858 | 1417 | 14 | -20, 40 | 17 | -19, 42 |
| AstraZeneca | 67 | 6 | 5858 | 1417 | 63 | 14, 84 | 64 | 16, 84 |
| Janssen (Johnson & Johnson) | 27 | 3 | 5858 | 1417 | 54 | -52, 86 | 54 | -54, 86 |
| Moderna | 11 | 1 | 5858 | 1417 | 64 | -191, 95 | 63 | -187, 95 |
| Pfizer | 24 | 3 | 5858 | 1417 | 48 | -172, 84 | 46 | -81, 84 |
| **Predominant variant during vaccination** |  |  |  |  |  |  |  |  |
| Alpha | 31 | 3 | 5858 | 1417 | 60 | -31, 88 | 64 | -19, 89 |
| Delta | 103 | 17 | 5858 | 1417 | 32 | -14, 59 | 34 | -12, 60 |
| Omicron | 172 | 30 | 5858 | 1417 | 18 | -7, 51 | 29 | -6, 52 |
| **Vaccine Effectiveness against admission with COVID-19 SARI** |  |  |  |  |  |  |  |  |
|  | Vaccinated patients <sup>b</sup><br>(n=186) |  | Unvaccinated patients<br>(n=3652) |  | COVID-19 Vaccine Effectiveness (VE) |  |  |  |
|  | Negative<br>(controls) | Positive<br>(cases) | Negative<br>(controls) | Positive<br>(cases) | Crude VE<br>% | 95% C. I | Adjusted<br>VE % | 95 % C. I |
| **Vaccination status** |  |  |  |  |  |  |  |  |
| Vaccinated | 156 | 30 | 2866 | 786 | 30 | -4, 53 | 26 | -11, 50.5 |
| **Vaccine Product (first dose)** |  |  |  |  |  |  |  |  |
| Unknown <sup>a</sup> | 94 | 22 | 2866 | 786 | 15 | -36, 47 | 8 | -47, 43 |
| AstraZeneca | 37 | 5 | 2866 | 786 | 51 | -26, 81 | 49 | -30, 80 |
| Janssen (Johnson & Johnson) | 8 | 1 | 2866 | 786 | 54 | -264, 94 | 40 | -386, 93 |
| Moderna | 8 | 1 | 2866 | 786 | 54 | -264,94 | 59 | -230, 95 |
| Pfizer | 9 | 1 | 2866 | 786 | 59 | -220, 95 | 61 | -211, 95 |
| **Predominant variant during vaccination** |  |  |  |  |  |  |  |  |
| Alpha | 18 | 3 | 2866 | 786 | 39 | -107 | 39 | -110, 82 |
| Delta | 66 | 7 | 2866 | 786 | 61 | 15 | 61 | 13, 82 |
| Omicron | 51 | 2 | 2866 | 786 | -1 | -67 | -11 | -84, 33 |

<sup>a</sup> These patients reported being vaccinated and had SMS confirmation of the same (with the date of vaccination) but their vaccination records were not verifiable on the national vaccine registry
<sup>b</sup> Number of vaccinated patients included in the estimation of VE against admission with COVID-19 SARI was reduced because of some vaccinated patients missing COVID-19 test results

Adjusted VE against admission to hospital with COVID-19 was not statistically significant overall (i.e., vaccinated versus unvaccinated) adjusted VE 26%, 95% CI -11,50.5. When stratified by period, VE against admission with COVID-19 was highest during the delta period (61%, 95% CI 13,82) (Table 3).

## Discussion

The value of surveillance systems during the COVID-19 pandemic cannot be overstated (9). Insights from these systems and other hospital-based studies have been useful in identifying those at increased risk for severe outcomes and informing both pharmaceutical and non-pharmaceutical interventions against the disease(6, 25, 26, 27, 28, 29). Our study found that patients admitted between May 2020 and December 2022 with SARI had a higher mortality rate, especially those with COVID-19, compared to their non-SARI counterparts. As in other studies, our study identified being advanced in age(5, 7, 10, 30, 31, 32, 33, 34), male(3, 5, 6, 7, 10, 32, 33), having leukocytosis(30, 32, 35, 36), anemia and/or a low oxygen saturation(34) at admission as risk factors for death in patients admitted with COVID-19 in Kenya. Similar findings have been reported elsewhere (5, 6, 7, 37).

Various contextual factors may explain the results presented. First, inadequate access to specialized care like invasive ventilation and disruptions in the supply of medical commodities may have prevented their use in patients who were at increased risk for poor outcomes. This may have contributed to the higher mortality observed among patients with SARI (38, 39). Secondly, variable testing and recording of co-infection with other infectious diseases like malaria and HIV which are endemic to our region limited the study’s ability to explore mortality among these subgroups. For example, routine testing for malaria was low with only 24.9% of adults tested. Unfortunately, routine testing for malaria was one of the services disrupted by the pandemic in Kenya and may have caused some of the patients with co-infection to miss out on treatment(40). While we report a reduction in hospitalisations for adults and in-hospital mortality during the COVID-19 period compared to the pre-COVID period elsewhere(18), our study still observed the disproportionate burden of mortality due to COVID-19 among the elderly and male patients compared to younger and female patients that has been reported globally.

Estimating vaccine effectiveness using observational studies, especially those in low- and middle-income settings such as ours, is challenging and not without shortcomings(41). We report a modest COVID-19 vaccine effectiveness of 34% against mortality due to SARI during the COVID-19 pandemic in Kenya. This VE appeared to be higher among patients who received the AstraZeneca vaccine product (64%). However, we could not definitively determine statistically significant differences in VE by vaccine products due to limited numbers of patients that received the different vaccines. Overall, these VE estimates are lower than those reported elsewhere in sub-Saharan Africa (42, 43, 44, 45, 46). Our estimates, while informative, should be interpreted with caution as they are reported against a backdrop of extensive community spread of COVID-19 before vaccine introduction(47), limited access to and availability of COVID-19 tests and low vaccine coverage with variable availability of the vaccine(14). For example, COVID-19 vaccine coverage among the study participants was 4% compared to a national vaccine coverage of 36%. Difficulty in data linkage across national, regional and local COVID-19 testing, vaccination registries and hospital admissions limited the patients included in these analyses. Consequently, the estimates reported are not without bias(48). Specifically, the effect of selection bias, healthy vaccinee bias, misclassification, waning immunity and differential depletion of susceptibles on the final estimates cannot be overlooked. Additionally, limited capacity to verify verbal vaccination histories and mixing of vaccine products due to variable availability of specific vaccine products hindered our study’s ability to estimate VE by all licensed vaccine products.

## Strengths and Limitations

A major strength of our study is the large sample size achieved during the study period. Where data on clinical outcomes in patients with COVID-19 in our region are limited(49), these results provide key insights on the pandemic within our setting. Our study developed a dynamic data collection tool that allowed real-time review of management practices and clinical outcomes with evolving knowledge on the disease. Whilst the study was not designed to evaluate emerging COVID-19 treatment modalities, its findings on the use of steroids in treating SARI have allowed for further studies to be conducted (50).

Despite including multiple sites spread across densely populated areas of the country and having a large sample, the results of this study may not be fully generalizable to the country. Our sample of COVID-19 positive patients represents only about 1% of all cases reported in the country. However, the results are still relevant both nationally and globally.

Relying on routine clinical data and documentation practices which differ by individual and by site limited the study’s ability to explore the effect of other known risk factors for death in COVID-19 such as multiple comorbidities at admission, social habits e.g., smoking, HIV infection or exposure to previous medication (6, 10, 29). While completion of demographic parameters was high, poor documentation practices and missing data for clinical parameters and investigations limited the number of patients included in the multivariable analysis. This variability in documentation may have introduced bias to our estimates. Additionally, variable documentation limited our capacity to describe the changing clinical pattern of COVID-19 by dominant SARS-CoV-2 variant or to explore the outcome in COVID-19 patients by evolving treatment practices.

Despite these limitations, the study identifies at-risk populations and highlights the need for improved documentation practices, access to specialized care and medical supplies, and more comprehensive testing for co-infections to improve the outcomes of COVID-19 patients in Kenya.

## Future of study

The COVID-19 pandemic has highlighted the need for effective hospital surveillance systems in Kenya(11). By monitoring disease activity within the population in real-time, using signs and symptoms, or preliminary diagnoses, hospital surveillance systems can indicate changes in disease trends that require immediate attention(51).

Standardization of the minimum clinical data documented at health facilities is necessary to effectively monitor disease activity. Developing a standardized admission tool or collecting key priority information from hospitals and transmitting these data to a national repository would enable data linkage and analysis, thus improving surveillance and planning. Additionally, expanding our study to include real-time on-demand analysis(10) could further enhance the effectiveness of the hospital surveillance system.

The future of VE studies nested upon observational studies in low- and middle-income countries like Kenya is bright. More efforts are increasingly being undertaken to optimise methodological approaches and build capacity to conduct these studies. Lessons from the COVID-19 pandemic will be key in developing key framework and designing longitudinal observational studies that can be rapidly adapted to conduct vaccine effectiveness studies in the setting of disease outbreaks.

## Conclusion

Our study has generated valuable clinical data on mortality among patients admitted to adult medical wards in the country and estimates of COVID-19 vaccine effectiveness during the COVID-19 pandemic. These data were useful in providing situational awareness during the first three years of the pandemic in Kenya and informing the national response measures. We also demonstrate the potential for routine clinical surveillance systems to inform public health activities in low resource settings.

## Data Availability

Data are available upon reasonable request

## Declarations Ethical approval

This study was approved by the Ministry of Health of Kenya (MOH), Scientific and Ethical Review Unit (SERU) of the Kenya Medical Research Institute (KEMRI/CGMR-C/203/4085), the Oxford Tropical Research Ethics Committee (OxTREC 44-20), and the London School of Hygiene and Tropical Medicine Research Ethics Committee (LEO 26950). Additional approval was sought from all participating hospitals and their respective county health departments. Individual consent for access to de-identified patient data was not required.

## Publishing

For the purpose of Open Access, the author has applied a CC-BY public copyright license to any accepted manuscript version arising from this submission.

## Acknowledgements

We are grateful to the hospital teams, including staff, patients and caregivers, and Ministry of Health colleagues, for supporting the Clinical Information Network and all clinical surveillance activities of the Kilifi HDSS. This article is published with the permission of the Director of the Kenya Medical Research Institute.

---

**Supplementary Figure 1:**
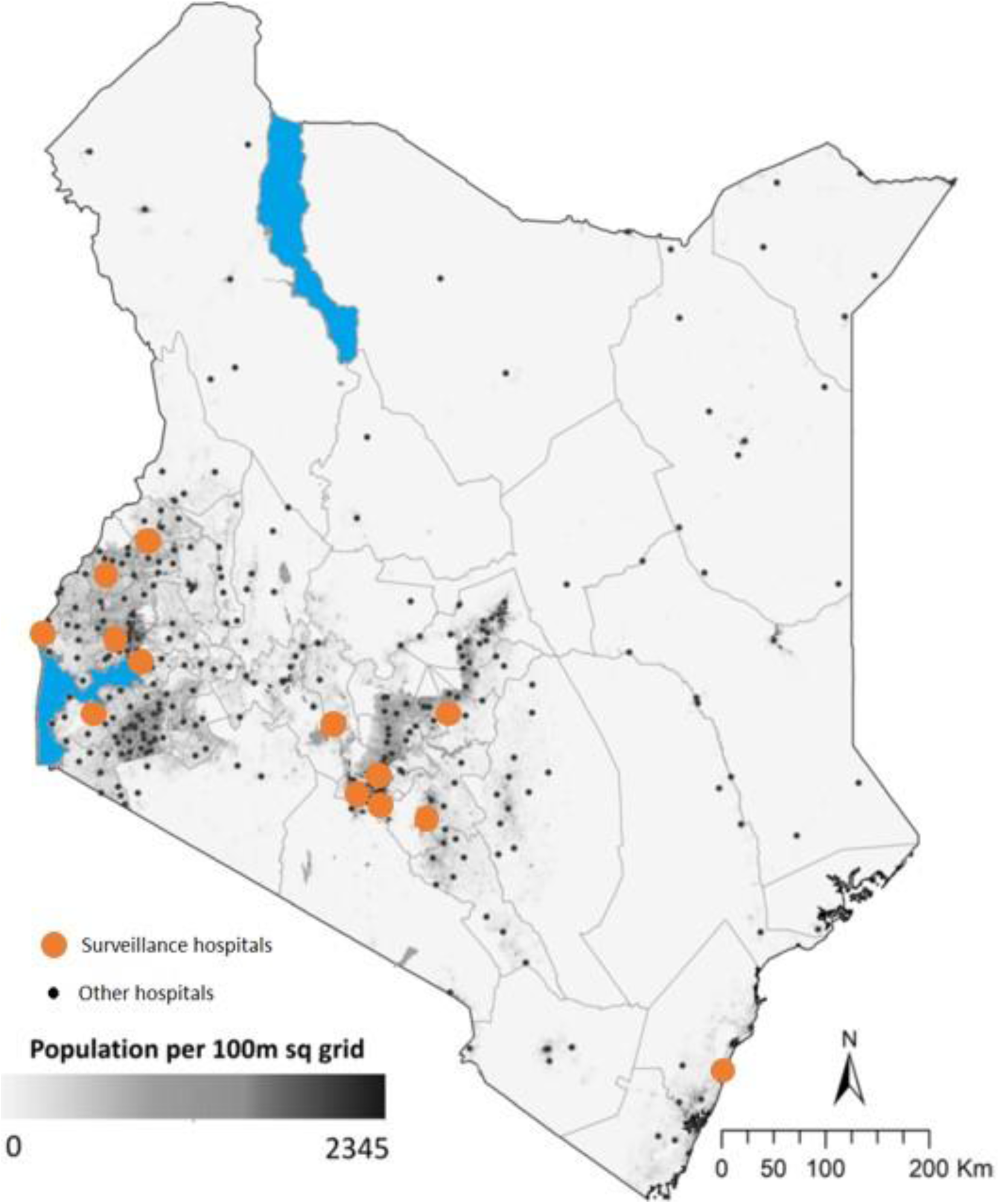
Surveillance study sites

**Supplementary Figure 2:**
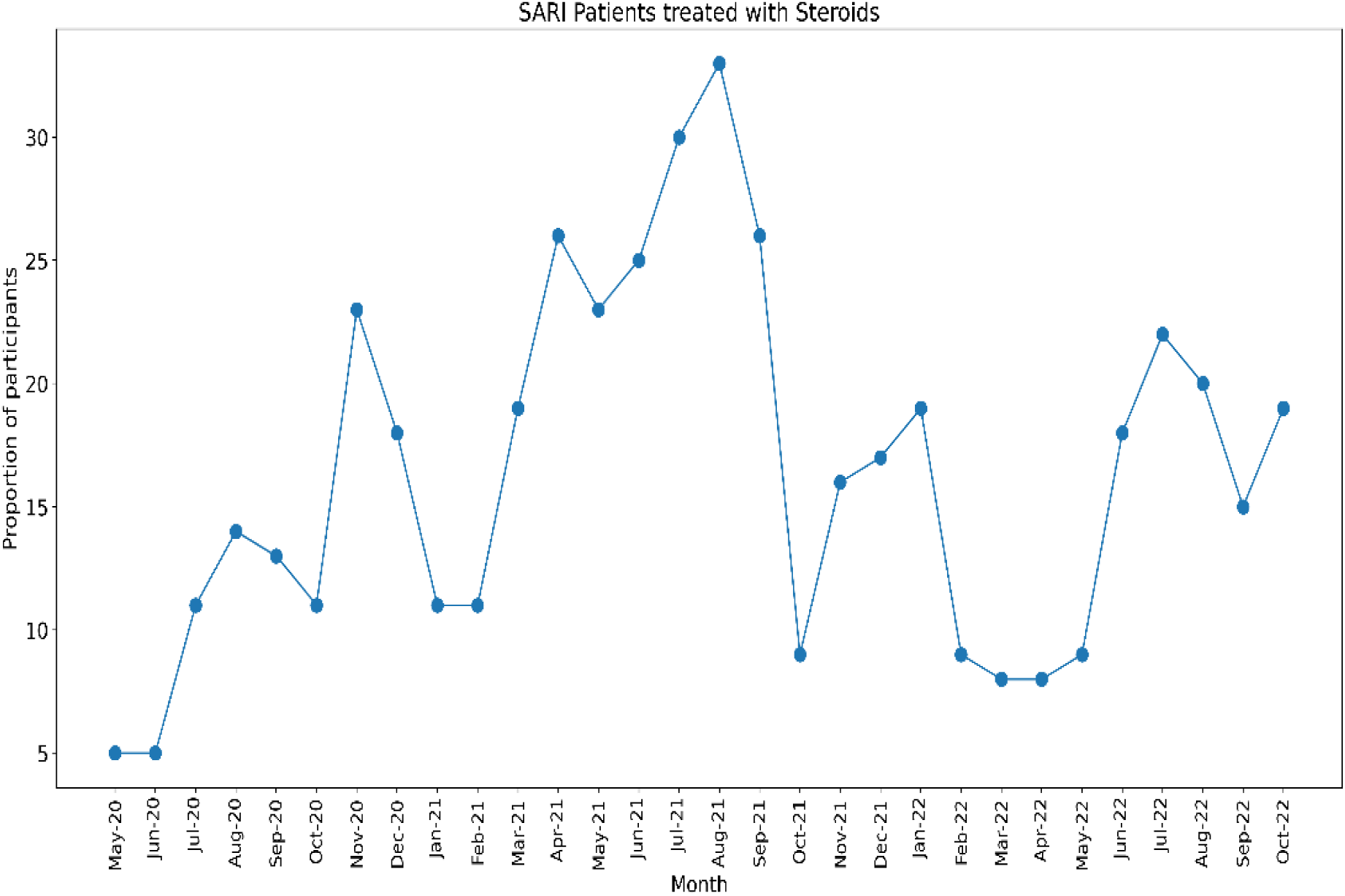
Proportion of Patients Treated with Steroids The figure below summarizes the proportion of severe acute respiratory illness (SARI) patients receiving steroids as part of their treatment regardless of their COVID-19 status. After the recommendation of steroid use in the treatment of COVID-19 was made by the world health organization (WHO) in May 2020, patients with SARI were increasingly managed with steroids regardless of their COVID-19 status

**Supplementary Table 1:** Distribution of SARI Patients by Year The table below summarizes the distribution of SARI patients’ outcome and management by year and quarter of the year during the study period

| Year | Quarter of Year | <sup>a</sup> Patient Characteristics |  |  |
| --- | --- | --- | --- | --- |
|  |  | % Deaths<br>(n = 8,848) | % Managed<br>in Isolation<br>(n = 2,835) | % Required<br>Specialised<br>Care<br>(n = 10,274) |
| 2020 | *May to June 2020 | 7.1 (627) | 4.3 (123) | 7.4 (756) |
|  | July to September 2020 | 11.9 (1,051) | 16.1 (455) | 12.2 (1,257) |
|  | October to December 2020 | 8.5 (753) | 20.5 (581) | 10.7 (1,097) |
| 2021 | January to March 2021 | 8.9 (788) | 12.5 (354) | 9.3 (953) |
|  | April to June 2021 | 12.6 (1,114) | 18.6 (527) | 13.1 (1,342) |
|  | July to September 2021 | 13.3 (1,176) | 18.2 (515) | 9.7 (991) |
|  | October to December 2021 | 7.0 (617) | 4.7 (133) | 5.3 (549) |
| 2022 | January to March 2022 | 8.0 (704) | 3.8 (108) | 11.1 (1,135) |
|  | April to June 2022 | 9.7 (861) | 0.1 (2) | 9.2 (943) |
|  | July to September 2022 | 10.2 (906) | 0.7 (20) | 9.4 (967) |
|  | October to December 2022 | 2.8 (251) | 0.6 (17) | 2.8 (284) |

<sup>a</sup>Chi square test of difference in distribution of patients by quarter in all three groups had a p value < 0.0001
\*Data for April 2020 not included in this analysis as the study had not begun

